# Environmental Mixtures Analysis (E-MIX) Workflow and Methods Repository

**DOI:** 10.1101/2024.12.20.24318087

**Authors:** Bonnie R. Joubert, Glenn Palmer, David Dunson, Marianthi-Anna Kioumourtzoglou, Brent A. Coull

## Abstract

Human exposure to complex, changing, and variably correlated mixtures of environmental chemicals has presented analytical challenges to epidemiologists and human health researchers. There have been a wide variety of recent advances in statistical methods for analyzing mixtures data, with most of these methods having open-source software for implementation. However, there is no one-size-fits-all method for analyzing mixtures data given the considerable heterogeneity in scientific focus and study design. For example, some methods focus on predicting the overall health effect of a mixture and others seek to disentangle main effects and pairwise interactions. Some methods are only appropriate for cross-sectional designs, while other methods can accommodate longitudinally measured exposures or outcomes. This article focuses on greatly simplifying the daunting task of identifying which methods are most suitable for a particular study design, data type, and scientific focus. With this goal in mind, we present an organized workflow for statistical analysis considerations in environmental mixtures data. This systematic strategy builds on epidemiological and statistical principles, considering specific nuances for the mixtures’ context. We also describe an accompanying online methods repository in development to increase awareness of and inform application of existing methods and new methods as they are developed and identify gaps in existing methods warranting further development.

## Introduction

Human exposure to complex mixtures of environmental chemicals has presented analytical challenges to epidemiologists and human health researchers more broadly. However, through recent advances from the statistical and quantitative communities, a wide variety of statistical methods for analyzing mixtures data now exist. In fact, in the last ten years, over 50 statistical methods for environmental mixtures analysis have been published, in part in response to the unmet needs noted in the 2015 National Institute of Environmental Health Sciences (NIEHS) workshop on statistical methods for mixtures ^1^, the 2017 NIEHS Powering Research through Innovative Methods for mixtures in Epidemiology (PRIME) program ^2^, and through investigator-initiated research projects supported by institutions or other US and international funding agencies. Most methods include open-source software for implementation, with accompanying vignettes or instructional documentation and simple example datasets. Although there remains a need for new methodology, the field has come a long way and now includes a rich collection of statistical methods available for analyzing mixture data from a variety of study designs (cross sectional, longitudinal exposures and/or outcome, time-to-event, etc.) and for a range of inferential interests (predicting overall health effect of the mixture, inferring which constituents significantly impact health, estimating main effects and interactions, etc.).

The collection of available methods and considerations for their applications has been described in several reviews^3–9^. To guide researchers navigating informed applications, methods are often organized into research questions. Braun et al. ^3^ present three key research questions, expanded to four by Hamra et al 2018^6^: (a) What is the effect of an aggregate mixture? (b) What is the effect of a sum of mixture components? (c) What are the independent effects of mixture components? and (d) What are the joint effects of mixture components? Gibson et al. 2019 ^5^ present a slight variation of these questions, tailored to an analysis dataset, reiterated by Joubert et al., 2022 ^9^ as methods designed to assess the: (a) overall effect estimation, (b) toxic agent identification (variable selection), (c) pattern identification, (d) a priori defined groups, and (e) interactions and non-linearities. Some reviews have also organized mixtures methods into two broad categories of supervised and unsupervised strategies ^5^, the latter focused on the exploration of patterns in exposure data, independent of a specific outcome or health endpoint. Although these guidelines enable methods categorization and understanding, it is common for one method to address more than one research question, and many methods may be appropriate in any given context. In addition to research questions and study design, researchers deciding between various methods also need to consider the distribution of the outcome and study design (continuous, binary, time-to-event, repeated measures etc.), the structure of the measured exposure data (e.g., varying in space and/or time), the size of the dataset, and other details.

Understandably, researchers may incorporate individual preferences in methods selection reflecting their training, recommendations from statistical collaborators, interpretability of the results, access to published methodology (open access publications), open access software, or other factors. Clear and freely available instructions ranging from minimal documentation accompanying *R* packages in *GitHub* to more detailed vignettes in CRAN or in eBooks, ^10^ and inclusion of methods in institutional coursework or short training programs such as the Columbia SHARP Mixtures Workshop ^11^ can ideally facilitate the new use and reuse of available methods. Nonetheless, researchers, particularly new researchers, may find it difficult to know where to start, which methods to select and why, and how many methods are applicable for any given analysis. As such, informed methods selection remains an issue for environmental health researchers when designing a study, secondary data analysis, dissertation or thesis project, grant application, or in response to peer review of a publication.

With these challenges in mind, we present the Environmental Mixtures Analysis (E-MIX) Workflow, to organize mixtures methods into meaningful categories that inform application. The E-MIX workflow builds on the available resources to date and covers key questions researchers should address when approaching a mixtures analysis. These questions are based on methodological principles of epidemiology, tailored to the context of environmental mixtures. In addition to informed application, this workflow can increase awareness of new methods. Importantly, instead of comparing methods in a competitive way, offering one as more or less preferable than another, E-MIX offers a neutral guide that facilitates leveraging multiple methods for a variety of contexts.

## Methods Considered

For the purpose of this seminar, we queried PubMed for statistical methods for environmental mixtures meeting the following criteria: 1) designed to examine the association between multiple (at least three) environmental exposure variables and a health outcome variable using data from human participants; 2) existing methods described in a literature review article on mixtures methods in the last five years (January 1, 2020 or later), a new method published in the last five years (January 1, 2020 or later), or a new method published in the last ten years (January 1, 2014 or later) that has been commonly applied in the mixtures epidemiology literature; 3) methods with findable open-source software such as an *R* package posted in *GitHub*; and 4) methods with clear and sufficiently detailed software documentation to inform application. For this manuscript, we present a subset of the identified methods where we could accurately present the method for each step in the workflow (Table 1). Additional methods will be available in the **NIEHS Environmental Mixtures Methods Repository**, described below.

**Table 1.**
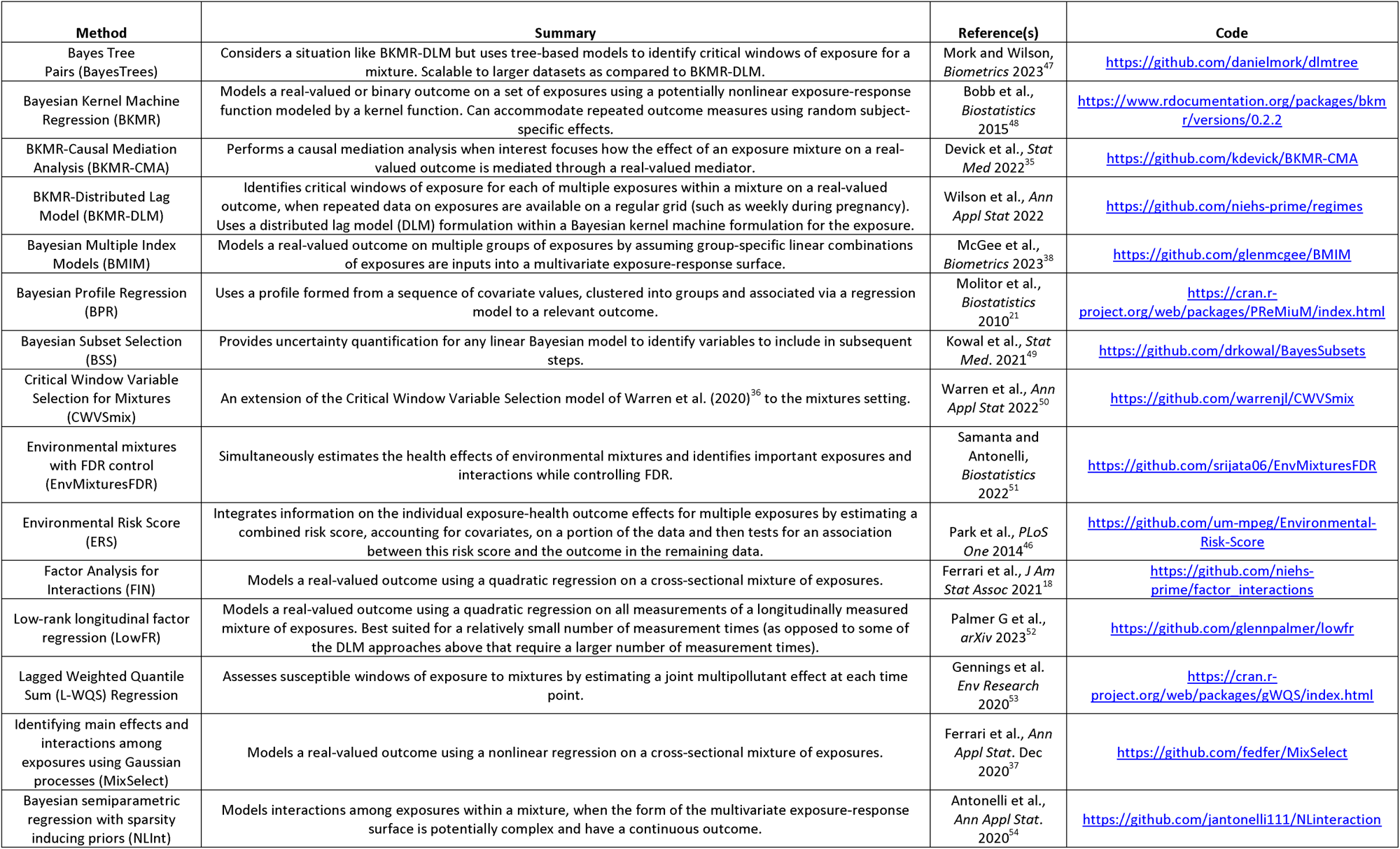
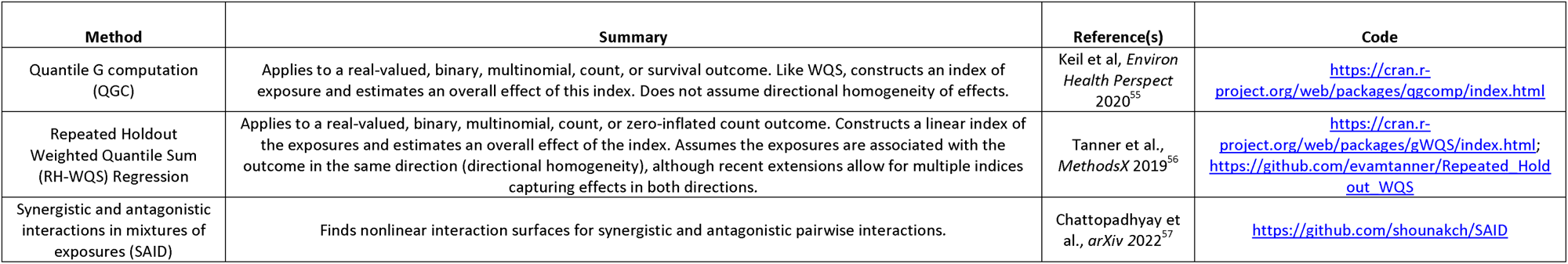
Mixtures Methods Considered.

## Environmental Mixtures Analysis (E-MIX) Workflow

The Environmental Mixtures Data Analysis (E-MIX) Workflow offers an organized series of steps for approaching environmental mixtures analysis in epidemiological data. The goal is to provide a guide for researchers having a particular dataset and inferential focus for their data analysis in hand. The steps in the workflow are intended to be followed sequentially, starting with the overall conceptual model for the analysis of interest (Step 1) and data processing and exploratory analyses (Step 2). Responses to prompts of the Steps 3 through 5 can be addressed in any order and will result in a list of relevant statistical models that can be applied to address a particular mixtures analysis context or hypothesis.

The final Step 6 includes considerations for model assessment and evaluation. The E-MIX Workflow is detailed below and summarized in Figure 1.

**Figure 1.**
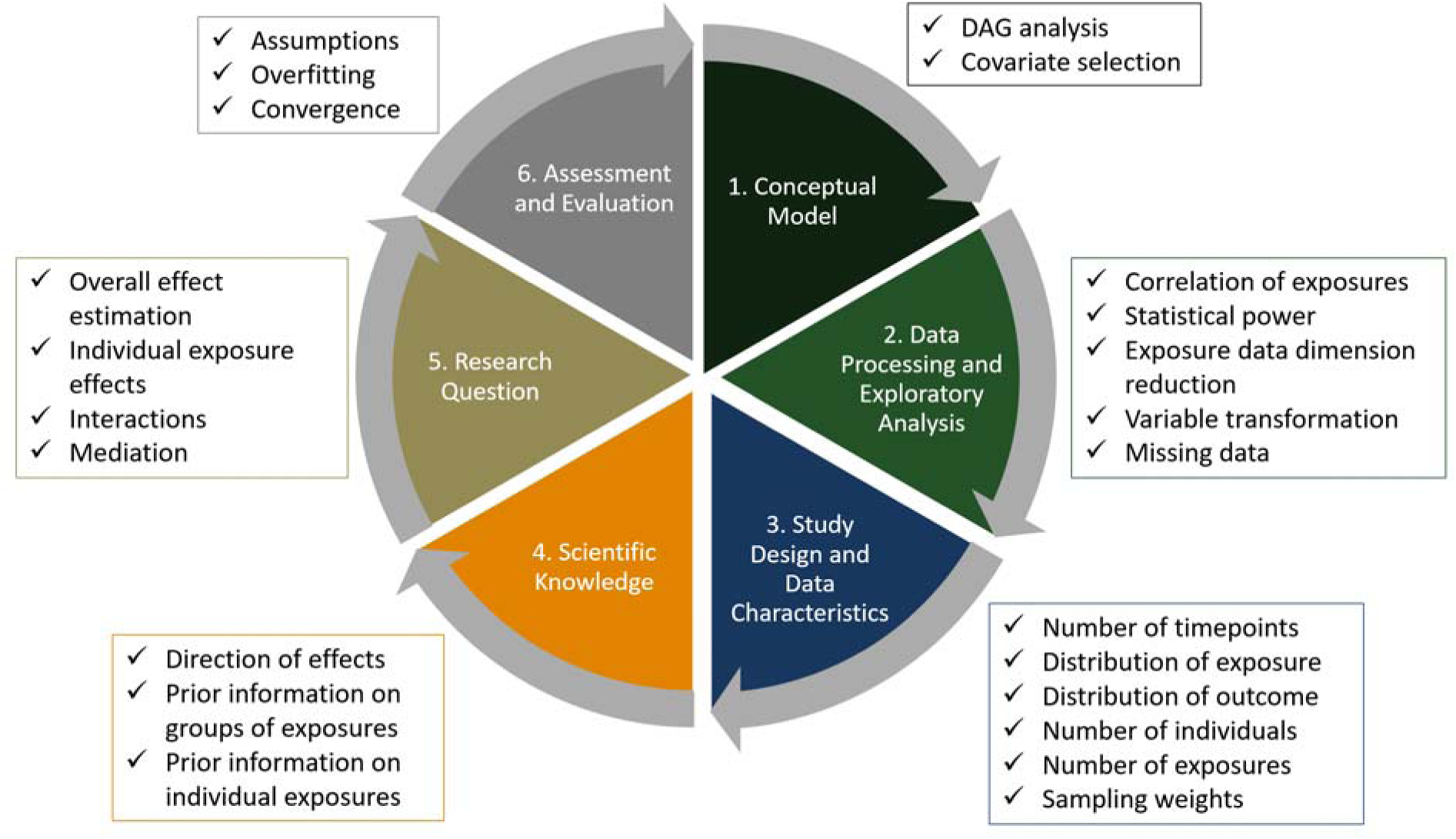
Summary of E-MIX Workflow Steps 1 – 6.

### Step 1. Conceptual model development

#### Directed Acyclic Graph Analysis and Covariate Selection

To hypothesize the causal relationships between the outcome and exposures considered to be part of the mixture, as well as covariates of interest, researchers can apply Directed Acyclic Graph (DAG) analysis. Examples of standard DAG analysis in epidemiology have been previously described (Lash et al., Modern Epidemiology, 4^th^ edition)^12^. Covariate selection should also be carefully considered, including whether a covariate is associated with both the exposure(s) and the outcome of interest, whether it operates as a modifier of the exposures-outcome association, or whether it should be considered as a mediator. For the mixtures context, Weisskopf et al. 2018^13^ describe example scenarios where including multiple correlated variables in a model such as a regression can increase (amplify) bias in the model, particularly when using variables representing biomarkers of environmental exposures. This bias is referred to as <u>co-exposure amplification bias,</u> and is important to consider in early stages of mixtures analysis. Overall, the conceptual model development phase should result in a list of variables of interest and an analysis dataset to apply to subsequent steps. In this design phase, researchers should also consider selection/collider bias.

### Step 2. Data Processing and Exploratory Analysis

#### Examine Correlation of Exposures

A key step in the analysis of environmental mixtures data is to explore the structure and magnitude of correlations among exposure variables. This exploratory analysis is helpful for several reasons. First, it can help inform which methods will be effective for a given dataset. For instance, if correlations among a set of exposures is 0.9 or above, it could be the exposures arise from the same source and almost always travel together, making it difficult for any mixture model to untangle the effects of individual exposures, especially if the sample size is small. In such cases, it may be more useful to employ careful dimension reduction prior to modeling the exposures or use a grouped variable selection method that quantifies the importance of the group of variables in the model. Knowledge of the magnitudes of the correlations can also help one understand what exposure scenarios, and therefore what contrasts of the mixture, are realistic. For instance, if two exposures have a pairwise correlation of 0.8, then it is unlikely that there are any subjects in the dataset having low levels of one exposure but high values for the other. In this case, predicting the mean outcome under a low-high exposure scenario would extrapolate outside of the observed data. If pairwise correlations are all low, there may not be many observations with extremely high levels of all exposures (i.e. all at 90% percentile).

#### Examine Statistical Power

Before performing analysis, it may be useful to investigate the probability that a given method will detect an association given an assumed data generation model, sample size, and effect size. This probability is referred to as statistical power and is particularly important in planning a study, as it can guide what sample size is needed, but can also be useful in selecting a model based on an assumed data generating mechanism and effect size. While closed-form formulas are available for power in very simple settings, complex data like the that in the mixtures setting generally require a simulation study. To facilitate this, the *mpower* R package implements simulation-based power estimation specifically for the mixtures setting ^14^.

#### Exposure data dimension reduction (optional)

It is possible, especially with highly correlated ambient exposures such as air pollution, that researchers may want to first identify patterns in the exposure space (e.g., air pollution sources) and reduce the dimension of the mixture prior to health analyses. Several methods can be used without including information about the health outcome (unsupervised methods). Example methods appropriate for mixture data include Principal Components Pursuit (PCP)^15^ or Bayesian Factor Analysis ^16^. This step is considered an optional step, depending on the exposure data characteristics. These methods can also be used for exposure pattern recognition, a mixtures-related research question that we do not cover in this manuscript.

#### Variable transformation (optional)

Another optional step prior to data analysis is to transform and scale the outcome, exposure, and/or covariate variables. It is important to standardize the exposure variables, so they are all on the same scale. Because some popular mixture models assume normality for a continuous outcome, it can be helpful to log-transform such outcomes if they exhibit a log-normal distribution (see Step 3, Distribution of the outcome). We consider these as standard data preparation steps, part of core graduate level curriculum for epidemiology and biostatistics. Some mixtures methods are based on a particular type of transformation of exposure data, such as weighted quantile sum (WQS) regression and quantile G-computation (QGC), which use quantiles of the exposure distributions by default. Example code for variable selection and transformation steps are available in the Columbia Mixtures Workshop GitHub (https://github.com/lizzyagibson/SHARP.Mixtures.Workshop).

#### Manage missing data

A critical step in data processing is to address missing data. Missing data can occur in several forms, such as information indicating an exposure is below the limit of detection (LOD) or an exposure value is simply not available. To our knowledge, only a few mixture methods formally incorporate missing values into a unified modeling framework. Herring (2010)^17^ proposed a nonparametric Bayes approach to modeling exposures and health outcomes when the exposures are subject to limits of detection. More recently, Ferrari and Dunson (2021)^18^ proposed Bayesian factor analysis for inference on interactions.

Because both the observed exposures and outcomes are jointly modeled by <u>latent factors</u>, this method can have missing exposure values as inputs into the method. Bayesian profile regression is another Bayesian model that jointly models the distributions of the exposure and outcome data, and therefore is able to accommodate exposure data below the LOD, although to the best of our knowledge has not yet been used for this purpose ^19–21^. Otherwise, most mixtures methods do not allow the inclusion of missing values, so rows with missing data must be removed, or missing data imputed. This typically applies to both supervised and unsupervised methods, although some unsupervised methods such as PCP^15^ can also handle missing data and values below the LOD.

If exposure data are subject to limits of detection, then the insights provided by Lubin et al. 2004^22^ for single exposure studies can be helpful as a general guide. Popular strategies such as replacing a value below the LOD divided by √2 can yield biased health effect estimates unless the percentage of observations below the LOD is small (5-10%). However, this becomes more complicated in mixture studies if different, say 10%, subsets of the data have values below LOD for different chemicals.

Alternatively, multiple imputation is an approach that generates multiple realizations of missing exposure data from an assumed model and then uses the multiple copies of the data in subsequent health effects analyses to account for the uncertainty induced by the missingness. The resulting multiple values of health effect estimates can then be combined to produce an overall effect estimate, and associated uncertainty, using rules developed by Rubin 1987^23^. Luben et al. 2004 described how this can be done using a log-normal model for an exposure distribution, and the experimental package *censlm* implements this approach (https://github.com/mikmart/censlm). One can also use multiple imputation to generate multiple realizations of exposure values missing more generally, rather than below the LOD. A popular approach for this task is Multiple Imputation by Chained Equations (MICE)^24^, with a robust R package for implementation^25^. In frequentist mixtures analyses, such as WQS regression or QGC, one can use MICE to obtain multiple realizations of missing values for multiple exposures, feed each resulting copy of the data into the mixture model, and combine the resulting effect estimates using Rubin’s rule. This approach assumes the data are “missing at random”; that is, the missingness mechanism depends solely on data that are observed, not on unobserved factors. While a formal, rigorous Bayesian approach would jointly fit the exposure imputation model and health model jointly, as in Herring 2010^17^, Ferrari and Dunson 2021^18^, and other work, Bauer et al. 2020^26^ approximated this approach by applying a Bayesian health model (BKMR, in this case) to each realization of data obtained by MICE and accounted for the resulting uncertainty by amalgamating the posterior samples from each run into a single posterior sample. Code for implementing this approach using BKMR is available at https://github.com/kdevick/bkmr_MI.

### Step 3. Study design and data characteristics

Following the conceptual design and data examination and processing steps, researchers can then address questions relevant to the epidemiologic study design and variable characteristics of their analysis dataset, as not all mixtures methods are suitable for all design/data contexts. Specifically, researchers should consider whether the design is longitudinal and includes measurements of exposures and/or outcomes at multiple timepoints and whether spatial data are examined. Researchers should also consider the distribution of the outcome and size of the dataset and may wish to run through these steps more than once, for different variable transformations (e.g., evaluating a continuous outcome in one analysis, dichotomous outcome in a separate analysis). Steps 3 – 6 are presented as yes/no (0/1) prompts for each method listed in Table 2 and can be addressed in any order.

**Table 2.**
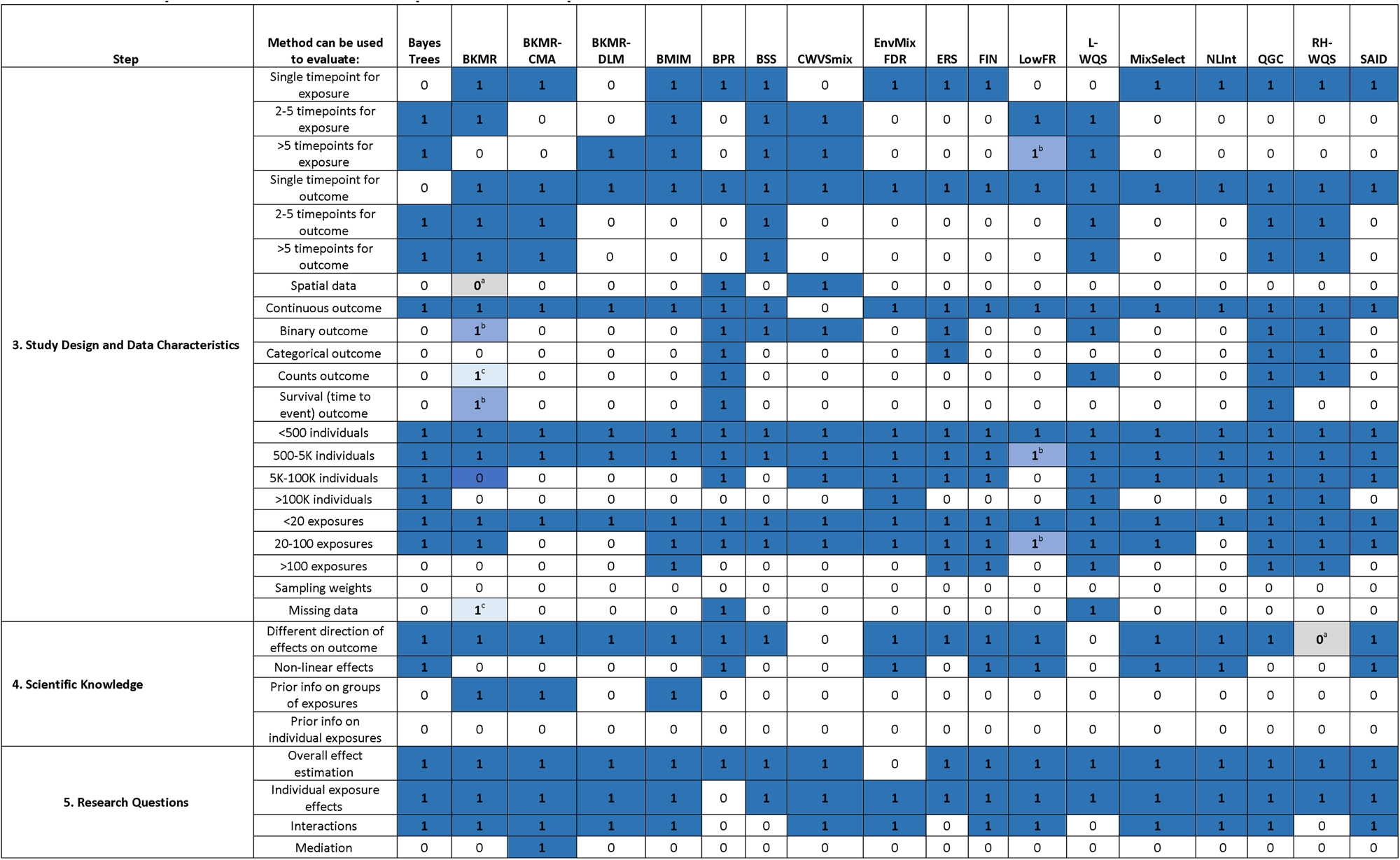
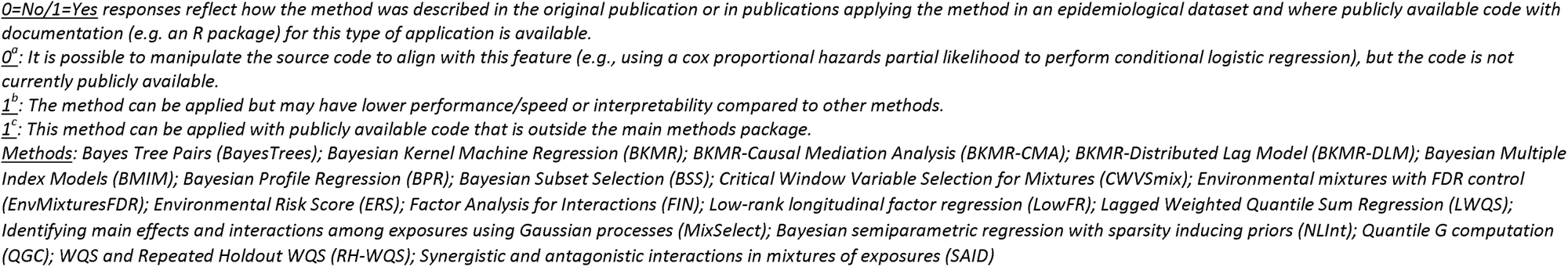
Summary of the E-MIX Workflow Steps 3 – 5 for Example Statistical Methods.

#### Single or repeated timepoints of exposures and outcomes

For this design/data characteristic, researchers can address **the question, Is the study longitudinal, with repeated measurements of exposures and/or outcomes or is there only a single timepoint for the measurement of exposure and outcome variables?** Historically, most methods for mixtures analyses have not considered multiple timepoints of exposure and outcome measurements. Now several models exist that can leverage longitudinal data. One can consider whether the analysis dataset includes repeated measurements of exposures and/or outcomes, and, if so, the number of timepoints of interest to include in a model. Often this requires considering a <u>distributed lag model</u> (DLM) or other methods that account for repeated timepoints. For example, CWVSmix, BKMR-DLM, Bayesian Tree Pairs, and Lagged WQS were designed to handle relatively many repeated, regularly measured values of multiple exposures (e.g., weekly measures of ambient air pollutants or temporally resolved tooth biomarkers of exposure). CWVSmix and BKMR-DLM assume smoothly varying effects of each chemical across pregnancy, so are limited to such designs, whereas Bayesian Tree Pairs, which uses tree-based methods to identify windows of susceptibility, could conceivably be applied to exposure data measured on fewer time points (e.g., trimester-specific measures). Development of methods that can address both big data and longitudinal data is one area that may warrant further research.

#### Spatial data

For this design/data characteristic, researchers can address the question, **Does the study include data with spatial variation and/or does spatial correlation among outcomes need to be considered in the model?** Studies have demonstrated the importance of fine particulate matter (PM_2.5_) on health outcomes, including cardiorespiratory and neurological outcomes, and that the effects can be spatially heterogenous ^27^. Importantly, the composition of particulate matter includes a mixture of chemicals including arsenic, sulfate, silicon, elemental carbon, and metals such as lead, iron, manganese, and zinc. As such, researchers working with air pollution data or other data that are spatially heterogenous may need mixtures models that can specifically address spatial heterogeneity. In addition to air pollution data, researchers may also wish to consider meteorological or geographical factors such as wind speed, ambient temperature, or humidity.

For mixtures datasets with spatial correlation in the exposure data varying in space and time such as air pollution composition data, researchers may also need to consider nonstationary processes to characterize space- and time-varying directional associations. Jin et al. ^28^ proposed such an approach, leveraging the role of wind direction on air pollution spread, while using Bayesian methods to allow for uncertainty in the directional DAG. Their “bag of DAGs” methodology speeds up computation for large spatiotemporal datasets relative to popular Gaussian process random effects approaches, while accommodating a more flexible covariance structure.

When there is residual spatial correlation in the outcome, ideally this would be modeled with spatially correlated random effects ^29^. While there are relatively few mixture methods that allow for spatial correlation among random effects, an exception is work by Mutiso et al. (2023)^30^ for small-area disease rates. This work modeled spatial correlation among areal disease counts using conditionally autoregressive random effects. Mutiso et al. indicate researchers interested in similar applications can contact the manuscript authors for example code.

#### Distribution of the outcome

For this design/data characteristic, researchers can address the question, **Does the dataset include a continuous, binary, categorical, count, or time-to-event (survival) outcome variable?** Some methods, such as BKMR, Bayes Trees Pairs, FIN, and MixSelect were developed for analysis using a continuous outcome variable. BKMR using a probit link for binary outcomes is also available. Other methods such as CWVSmix are suitable for binary outcomes but not continuous, categorical, counts, or survival outcomes, whereas QGC and WQS are appropriate for binary, categorical, and continuous outcomes. QGC can also handle survival (time to event) outcomes. For models that assume normality for the residuals of a model using a continuous outcome, it can be helpful to log-transform such outcomes to improve the normality assumption for the model errors.

#### Size of the dataset

The size of a dataset is commonly described as the number of individuals and the number of variables/exposures of interest to include in a specific model. Thus, for this design/data characteristic, researchers can address the question, **How many individuals and how many exposures/variables are included in the dataset for analysis?** For extremely large datasets, such as administrative data, applying a mixtures model that is not suitable for big data can result in extensive computational time and possibly issues with model convergence. To identify the best method, researchers can consider whether an analysis includes less than 500, 500-5,000, 5,000-100,000, or over 100,000 individuals and whether the dataset includes less than 20, between 20 and 100, or over 100 exposure variables. While multiple methods considered in this workflow handle settings with over 100 variables, we do not consider the broader set of methods designed to analyze data on the exposome, defined as encompassing an individual’s life-course environmental exposures, as there are related reviews available elsewhere ^31,32^. Because many exposomic studies focus on 100s to 1000s of biomarkers of exposure, a popular approach to analysis is an Exposure Wide Association Study (ExWAS), the most common form of which is a discovery-based approach that seeks to identify important phenotype-exposure pairs, while controlling for multiple testing^31^. In contrast, the mixture methods considered in this workflow focus on one or more of the research questions outlined in Step 5 (*Research Questions*).

#### Survey or sampling weights

For this design/data characteristic, researchers can address the question, **Are there survey or sampling weights to include in the analysis?** Many researchers leverage large survey data such as data from the National Health and Nutrition Examination Survey (NHANES). Traditional epidemiological models using these data can apply weights to account for oversampling strategies. Few mixtures methods can incorporate these weights, which is a known limitation for the field. However, anticipating how methods expand and evolve over time, we include this component in the workflow.

### Step 4. Scientific Knowledge

It can be helpful to utilize existing information from the literature, preliminary data, and/or toxicology in a model. For example, researchers can consider whether the exposure-outcome association (effect) of each component in a mixture is expected to operate in the same direction on the health outcome, or whether the model should allow effects in different directions. The researcher may also know that the exposure-response relationship is likely to be non-linear. There may be known structure or *a priori* information about groups of the exposures, such as chemical groups, that should be included in the statistical model. Some methods can leverage this information and group exposures in the modeling (e.g., BKMR hierarchical grouping extension, BMIM). Researchers may also wish to include individual information about exposures in the model, such as biological, toxicological, or other chemical features. For example, BMIM can incorporate the toxic equivalency factor, or related features, of subsets of exposures^33^. To consider this scientific information, researchers can consider the following questions: **Are exposures hypothesized to act in the same direction, or should the model allow for effects to operate in different directions? Are the exposure-response relationships likely to be non-linear? Is there biological, toxicological, or other information about the potential effects of the exposures such as chemical groups that should be included in the statistical model? Are there chemical properties/features to include in the model?** Responses to these prompts can be particularly critical to identifying relevant methods, as many existing methods are not able to address all these contexts.

### Step 5. Research Question

For this step, researchers can address the question, **What is the research question of interest for this analysis?** Identifying the research question and the overall goal of the analysis is critical to mixtures method selection. Although these have been described a few different ways in the literature, questions can roughly align with the following. Of note, these questions are specific to supervised strategies (including both exposure and outcome data). Unsupervised analyses can be considered earlier in the workflow (Step 2).

#### Overall effect estimation

Researchers often wish to determine the overall or aggregate effect of the mixture of exposures on a health outcome. Some interpret this to represent the effect of a sum of mixture components (separately distinguished in Hamra et a. 2018). Most presented methods can be applied to this research question; however, it is worth noting that the definition of “overall effect” varies from method to method. That is, the exposure contrast to which the overall effect corresponds is typically methods specific. Often, a method is selected not for addressing this research question alone, but for being able to address this question as well as one or more other research questions.

#### Individual exposure effects

Researchers are also often interested in the independent effects of mixture components. This has also been referred to as toxic agent identification or variable selection in Gibson et al. 2019 and Joubert et al. 2022 and can identify the “toxic agents” or “bad actors” in the mixture of chemical exposures. Most methods presented here can also address this research question.

#### Interactions

Examining the joint effects of mixture components or potential interactions is another common goal. Most methods can accommodate user-defined interactions by including products between two exposures as a new mixture. Among the presented methods here, most methods are able to address interactions when examining a single timepoint of exposure and outcome. However, fewer methods can address the effects of interactions within a mixture at multiple timepoints. Potential strategies include Bayes Tree Pairs, BKMR-DLM, CWVSmix, LowFR, MixSelect, and SAID (Table 2).

#### Mediation

Examining the effect of a mixture that mediates the association between an exposure and an outcome is a challenging area of mixtures. Bellavia et al., 2019 ^34^ describe strategies for approaching mediation analysis with environmental mixtures, including multiple regression, reducing the mixture to a single mediator, reducing the number of mediators, hierarchical modeling, and a two-stage approach by using a mixture method to select specific mediators. Devick et al., 2019^35^ present the use of BKMR-Causal Mediation Analysis (BKMR-CMA) for examining the effect of a mixture as an exposure on a health outcome, considering a single mediator. Because this method includes publicly available R code for implementation, we include it in the methods presented in Table 2. Further work with publicly available software for examining the effect of mixtures as the mediator between an exposure and outcome of interest, with publicly available software for implementation, is needed.

### Step 6. Assessment and Evaluation

After considering the above questions, a list of available methods for each research question can be obtained, with context and rationale for these recommendations. It is important to examine results with scrutiny, particularly given the complexities and nuances of mixtures models. Some example assessment considerations are noted here but should not be considered exhaustive.

#### Assumptions

It is important to examine important assumptions of each model identified relevant for a specific scenario. This is best considered prior to selecting and implementing a particular method. Many of the methods assume residuals in a model for continuous outcomes that are normally distributed with constant variance. Other models may have more specific assumptions. For example, LowFR assumes the exposures can be modeled with a multivariate normal distribution, and the expected outcome is well-modeled as a quadratic function of the exposures. Assumptions for each model should be carefully reviewed in model selection and interpretation of results.

#### Overfitting

Some methods have model-specific assumptions that should be tested, and each model should also be assessed for model fit and performance. For example, a model may appear to fit well to the data used to fit the model, but particularly for highly flexible models, this could simply be a result of interpolation for those specific observations, rather than learning a true underlying relationship. To guard against this, models can be evaluated on “out-of-sample” data. That is, researchers can divide the full dataset into non-overlapping “training data” and “test data” sets, fit the model to only the training set, and then evaluate the quality of predictions of that fitted model on the test data. This can be repeated multiple times with different training/test splits, in a process called cross-validation. The process of evaluating predictions on data not used to fit the model is commonly referred to as evaluating <u>out-of-sample-predictive performance</u>, and strong performance in this evaluation suggests greater confidence in the inferential conclusions generated by the model.

#### Convergence

Several existing mixture methods employ Bayesian methods for modeling fitting and inference. The Bayesian framework characterizes uncertainty in model parameters by conceptualizing them as random variables, and inference proceeds by summarizing a probability distribution, known as the posterior distribution, reflecting what values are more or less plausible, given the data. For many models, the posterior distribution of the model parameters is often not available in closed form, but samples from it can be generated using a stochastic Markov Chain process. This process, known as Markov Chain Monte Carlo (MCMC), typically takes multiple sequential samples to reach equilibrium and sample from the actual posterior distribution, also known as <u>MCMC convergence</u>. It is essential in applied Bayesian statistics to diagnose whether an MCMC algorithm has converged, so that one is confident that the resulting point estimates, credible intervals, and other summary statistics accurately characterize the posterior distribution of the model parameters.

There are multiple options for diagnosing convergence in available Bayesian models for environmental mixtures. A popular approach is to visually assess a plot of the sequential generated posterior samples (trace plot) and assess if these values randomly vary around a stationary mean. One can generate these plots manually using the returned posterior samples. Some software includes functions to generate these plots automatically and make available tutorials demonstrating how to interpret the resulting plots. See https://jenfb.github.io/bkmr/overview.html for BKMR. Another approach is to run multiple Markov Chains starting from different initial values and assess whether the resulting posterior samples appear to have been generated from the same posterior distribution. The R package coda can be used for analyzing MCMC output from any model to diagnose convergence. Others have developed packages that provide the ability to similarly run multiple chains and diagnose convergence for specific mixture models, such as *bkmrhat* for BKMR models. Regardless of the code applied to diagnose convergence for a given MCMC sampler, this should be standard practice when applying Bayesian models for environmental mixtures data.

## Environmental Mixtures Methods Repository

The workflow presented in this manuscript can be considered a stand-alone guide. It offers a systematic strategy for approaching mixtures analysis and can be applied to a wide range of mixtures methods, not just those presented tables and text of the manuscript. However, it is expected that new methods and modifications to existing methods will develop over time. As such, we present the **NIEHS Environmental Mixtures (E-MIX) Methods Repository**, currently in development. The E-MIX Repository is a NIEHS-managed web tool, where researchers can implement specific steps in the workflow and/or obtain more information about available mixtures methods (Supplementary Files 1 and 2). The E-MIX Repository follows the layout of other tools available on the NIEHS website such as the NIEHS Epidemiology Cohorts Faceted Search Tool (https://tools.niehs.nih.gov/cohorts/) or the Disaster Research Response Resources Portal (https://tools.niehs.nih.gov/dr2/). Following review of the overall purpose and workflow (Supplementary File 1, page 1), researchers interested in identifying an appropriate mixtures method for a given analysis can complete a faceted search (Supplementary File 1, “Describe your Data,” pages 2-4). This search provides results showing relevant methods for a given context (Supplementary File 1, “Search Results,” page 5). Researchers can then review details for each method as well as the original method paper(s), available software for implementation, and contact(s) for inquiries (Supplementary File 1, “Details Example,” page 6). Example searches are also available on the landing page (Supplementary File 1, “Example Searches”, bottom of page 1). The repository will enable a sustainable and neutral resource for sharing information about mixtures methods and provide a starting place for researchers approaching mixtures analysis. Methodologists will also be able to submit new methods to include in the repository (e.g., with information aligned to characteristics presented in Table 2) or to edit details of methods for which they serve as the contact. Close engagement between NIEHS web and program staff and method authors/contacts will ensure the presented material is accurate and up to date.

As the online repository develops, an alternative strategy for identifying methods is feasible by using the accompanying excel file (Supplementary File 2). The first row of this file can be completed by a researcher with an example dataset in mind (by entering “0” or “1” for each cell). One can then filter key columns to just results of “1.” This will reduce the full list of methods to a smaller set with matching characteristics to the dataset/application of interest. Researchers can then review details presented in Table 1 for the subset of methods, including links to R code for application and the original method publication(s).

### Example Applications

To describe example applications to the E-MIX workflow, we considered two publicly available datasets from the cross-sectional 2001-2002 National Health and Nutrition Examination Survey (NHANES) and the longitudinal Follow up to the Childhood Autism Risks from Genes and Environment (CHARGE) Study (ReCHARGE). The NHANES dataset was previously described and used by Mitro et al.^36^ to examine the association between exposure to persistent organic pollutants (POPs) and leukocyte telomere length (LTL). This dataset has been used extensively for training including the Columbia Sharp Mixtures Workshop ^5^, as well as for demonstrating applications of new methods such as FIN^37^ and BMIM^38^.The original dataset includes 1,330 adults, 18 exposures from three chemical groups, a continuous leukocyte telomere length outcome, and both continuous and categorical covariates. For the example application to the E-MIX workflow, since most of the methods we investigated do not automatically handle missing data, we considered a complete-case analysis (i.e., removed missing data), reducing the sample size to 1,003. However, note that any of the methods could be applied in combination with the multiple imputation approach described above in step 2 of the workflow. The telomere length variable is natural log-transformed and scaled to better follow a normal distribution, and chemicals are moderately to strongly correlated, particularly within each of the three chemical groups (see Figure 1 from Gibson et al., 2019 ^5^). Several covariates are available for inclusion in the model, as previously described ^5,36^.

The ReCHARGE study is a case-control study of 884 children ages 2-5 years with autism spectrum disorder (ASD), non-autistic developmental delay, or typical development population controls ^39–41^. Chemical measurements for this example dataset were measured by the Human Health Exposure Analysis Resource (HHEAR) data repository ^42^. This resulted in data for 83 chemicals across five groups (trace elements, pesticides, phthalates, phenols, and perfluorinated chemicals) in urine and plasma collected from children as well as information on education, race/ethnicity, sex, and maternal smoking during pregnancy and through follow up. Removing missing data reduces the analysis dataset to 601 individuals with ASD phenotypes, 62 chemicals, and covariates. The continuous cognitive score outcomes did not follow a normal distribution and the chemicals were moderately to weakly correlated across and within chemical groups (see Bennett et al., 2022 Supplementary Figures 1 and 2 ^39^).

For the example applications we first assigned a value of 0 (“No”) or 1 (“Yes”) for each prompt in the E-MIX workflow, steps 3 - 5 (Table 3). The NHANES application considers a single timepoint of exposure, single timepoint of the outcome, continuous outcome, fewer than 5,000 individuals, fewer than 20 exposures, no requirements on the directions of effect, and no prior information on groups of exposures. For the research question addressing overall effect estimation, relevant methods identified included BKMR, BKMR-CMA, BMIM, BPR, BSS, ERS, FIN, MixSelect, NLInt, QGC, RH-WQS, and SAID (Supplementary Table 1). Because this list includes 12 methods, researchers may wish to reduce it further by determining which methods are also applicable to other research questions of interest and/or whether some methods are not needed for a given context. For example, because BKMR-CMA is designed for examining mediation, a researcher may wish to remove this method from the list if mediation analysis is not the goal. Similarly, if interactions are not of primary interest, FIN could be removed or considered in a later analysis. In general, retaining a larger list will allow the researcher to consider methods with similar overall goals/parameters but that may vary in performance or offer different advantages for a particular application (e.g. computational time, model assumptions, model fit, etc.). However, if the researcher has stricter goals in mind such as including prior information on the groups of exposures and/or examining different directions of effects, they may prefer to start with a smaller list (in this example, BKMR, BKMR-CMA, and BMIM). An advantage of the E-MIX Workflow is that a researcher can try various combinations of responses to workflow prompts to explore what methods may work for their data and inference goals and apply multiple models. Importantly, after identifying a list of relevant methods, research must still scrutinize the assumptions and evaluation details of each method, as described in Step 6.

**Table 3.**
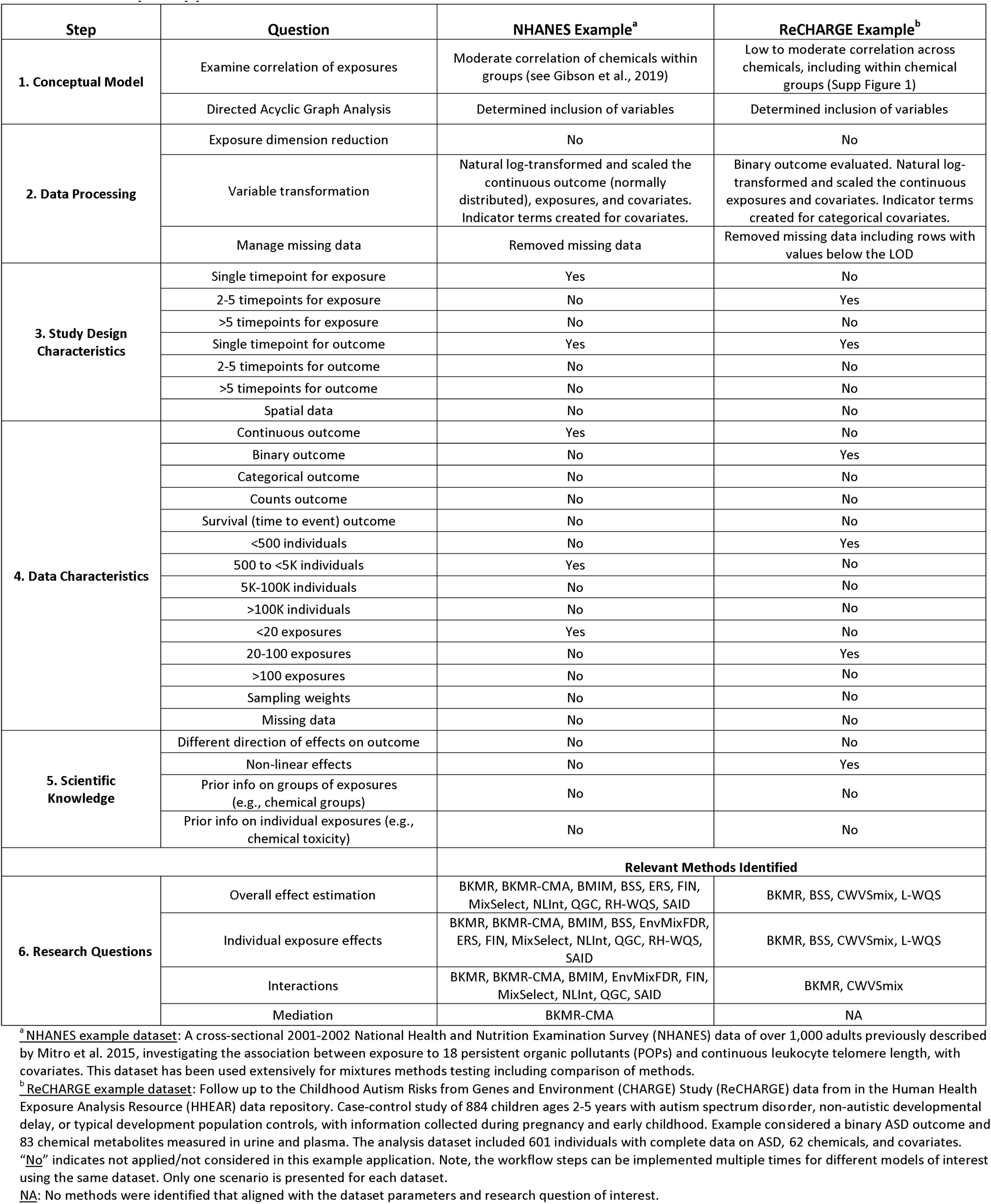
Example Applications of the E-MIX Workflow.

The ReCHARGE example application considers 2-5 timepoints of the exposures, a single timepoint of the outcome, a binary outcome, fewer than 500 individuals, fewer than 100 exposures, no requirements on the directions of effect, and no prior information on groups of exposures (Table 3). For the research question addressing the individual exposure effects, BKMR, BSS, CWVSmix, and L-WQS were identified (Supplementary Table 2). As previously noted, researchers can try different combinations appropriate for this dataset and model of interest to identify relevant methods for further scrutiny.

## Discussion

Informed statistical analysis of environmental mixtures data is a topic extensively explored in reviews, workshops, and short courses. These remain excellent guides, particularly for researchers new to the mixtures research space. The E-MIX Workflow expands these resources by presenting a systematic strategy to approach mixtures analysis, integrating concepts from epidemiological methods, practical considerations of the datasets, scientific knowledge about the data, research questions of interest, and statistical considerations for model assessment and evaluation. Prior resources have not offered a formalized integration of epidemiological and statistical concepts for mixtures in this way. The online E-MIX methods repository provides a unique space for sustainably sharing and updating available methods and for implementing the workflow.

It is important to note that several methods may be equally appropriate for a specific context. For example, researchers may follow the workflow steps and identify four to five or more methods to consider. We do not present a comparison or contrast of methods or recommend one method over another based on a metric of performance. Rather, the E-MIX Workflow can be used to identify several appropriate methods for a specific scenario to educate researchers (particularly new researchers or trainees) and inform application. Subjective decision making will still be required, but the assessment and evaluation steps are critical. We also encourage researchers to consider presenting results for more than one or two methods, thus evaluating sensitivity of the results.

In this workflow we have focused on mixture analyses differentiating between scenarios with less than 20, between 20 and 100, and over 100 exposure variables. While multiple methods considered in this workflow handle settings with over 100 variables, we do not consider the broader set of methods designed to analyze data on the exposome or other -omics data, as there are reviews of this field available elsewhere ^31,32^. Similarly, we do not consider important components of complex data analysis, including pre-processing steps for larger exposomic data analysis ^43^ or the analysis if multi-omics data ^44^. We encourage readers to also consult these and related methods for guidance on methods most suitable for large-scale ‘omic data.

The E-Mix workflow considers whether a method can handle missing data or requires complete case analysis. This can be an important limitation for many contexts, where the requirement for complete case analysis can substantially reduce the final sample size. This emphasizes the importance of the first few steps considering thoughtful variable selection. Of note, the strategies themselves for performing imputation were not assessed in this paper as they were considered outside the current scope. We also acknowledge that for many methods, users interested in modifying the original source code can include modifications for handling missing data. For this paper, we only considered what is currently presented in publicly available code. Other instances where source code and data structure can be modified to accommodate a desired feature include conditional logistic regression in some methods such as QGC using a Cox partial likelihood (https://github.com/alexpkeil1/qgcomp/). BKMR has also been modified for application to count data, accounting for spatiotemporal heterogeneity ^30^. Further, while the original implementation of WQS leveraged the assumption of directional homogeneity, recent work has sought to modify this assumption by incorporating two indices to allow associations to examine both directions of effects ^45^.

Researcher preferences for <u>index models</u> or <u>response surface models</u> are not fully incorporated in the workflow but should be considered carefully when identifying suitable methods for a given analysis. Both have distinct advantages and disadvantages, and in some ways are complementary. Linear index models reduce the dimension of the mixture into a small number, often only a single, exposure summary, which makes interpretation much more straightforward. However, these indices are often but not always formed making strong assumptions, for example assuming linearity and additivity of effects. If these assumptions hold, these models typically provide higher statistical power to detect effects, due to model parsimony. However, if these assumptions do not hold, inference can be biased due to model misspecification. Response surfaces, in contrast, are usually modeled flexibly, which allows for the estimation of nonlinear and non-additive effects and decreases the risk of model misspecification. However, this flexibility requires more data, especially in higher exposure dimensions, and can have lower power than index methods when the simpler assumptions hold or nearly hold.

We characterize whether the presented methods can include weights, such as survey or oversampling weights. However, methods taking weights into account are not well represented in the mixtures literature and warrant further development. The workflow also does not consider matched study designs, retrospective case-control studies, prediction analyses as opposed to association analyses, and causal inference, or intervention studies. Future iterations of the workflow could be produced to include more methods appropriate for these designs. We also did not present methods specific to modification by factors that are not members of the mixture. This can theoretically be addressed by any of the methods through stratification or sub-setting the data if the number of subgroups is small, although in certain scenarios there may be more statistically efficient methods that pool some information across groups (e.g. interaction analyses that assume the effects of exposures vary by subgroup, but the effects of confounders do not vary by subgroup).

We also note the potential to apply two mixtures methods sequentially, particularly unsupervised and supervised strategies. This can be a helpful way to approach analysis of high dimensional datasets when interested in applying a method not well suited for big data. For example, in step 2 of the workflow, an exposure data dimension reduction step can be applied to reduce a high number of exposure variables of interest to a smaller set of clusters or components, especially if interest lies in exposure pattern recognition. Then researchers may leverage methods identified in Step 3 suitable for datasets with 2 – 5 exposure patterns. Researchers must also consider the model assumptions of both methods applied. Another example is the use of penalized likelihood, particularly the elastic net, to form an exposure index, known as an environmental risk score, on a portion of the data, that can then be tested in the remaining data^46^.

A unique strength of this work is the development of the E-MIX public repository of mixtures methods, which will evolve over time. The repository will also highlight important research gaps in existing mixtures methods and/or the need for potential extensions of approaches to include other complex data such as larger scale exposomics or multi-omics. Additional training resources and methods development to address precision environmental health, causal methodology, risk assessment, and mediation approaches are important areas warranting further research and integration in this resource.

## Data Availability

The original NHANES data used in this exercise and example code for data formatting and some methods is available in the supplemental material of Gibson et al., 2019 (https://github.com/lizzyagibson/Mixtures.Workshop.2018). The original RECHARGE data was obtained from the publicly available data in the Human Health Exposure Resource (HHEAR) Data Repository under CHEAR project #2016-1461; doi 10.36043/1461_222, 10.36043/1461_219, 10.36043/1461_630_2022.2. from the HHEAR Data Center: https://hheardatacenter.mssm.edu/.

https://github.com/lizzyagibson/Mixtures.Workshop.2018

---

**Supplementary Table 1.**
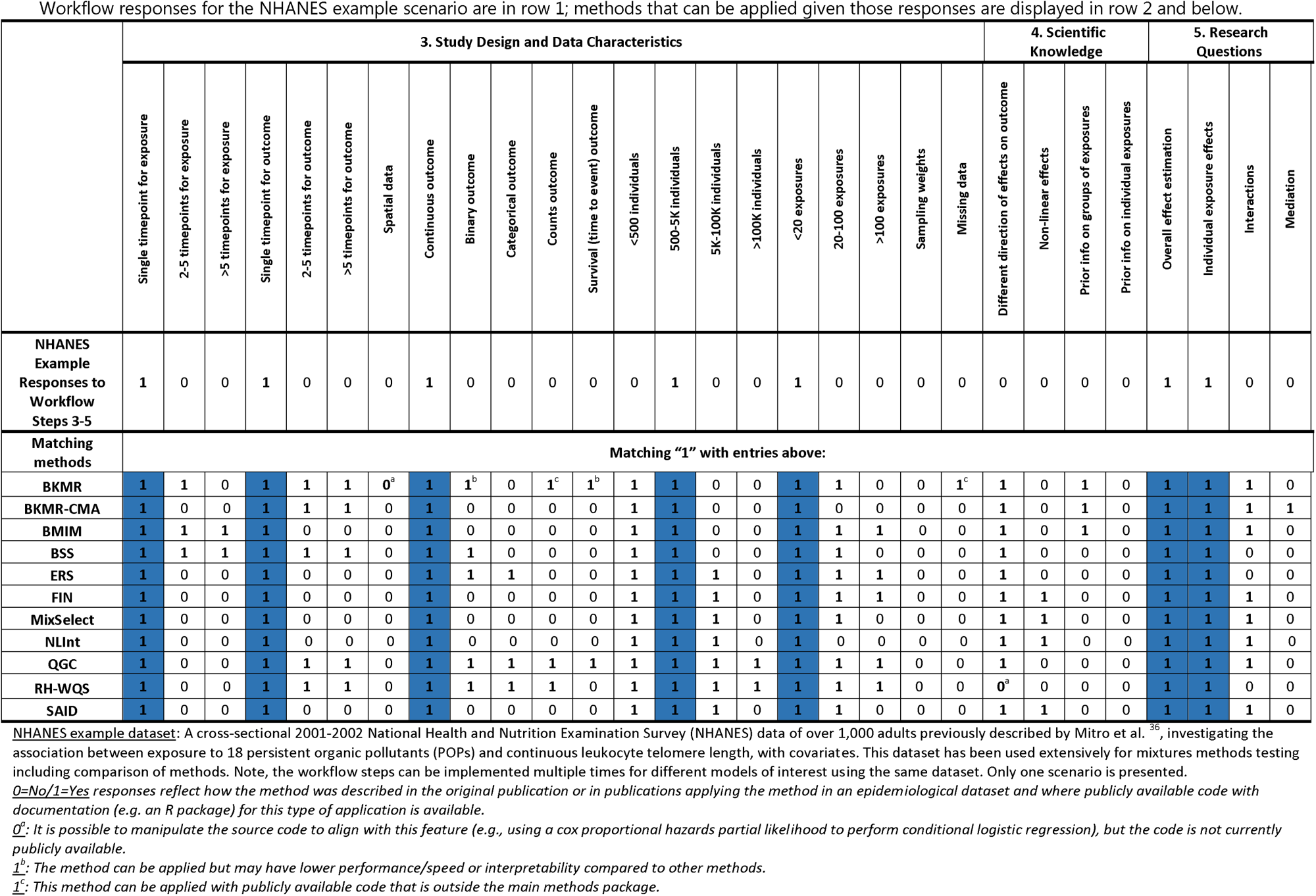
Application of the E-MIX Workflow Steps 3-5: NHANES Example Data.

**Supplementary Table 2.**
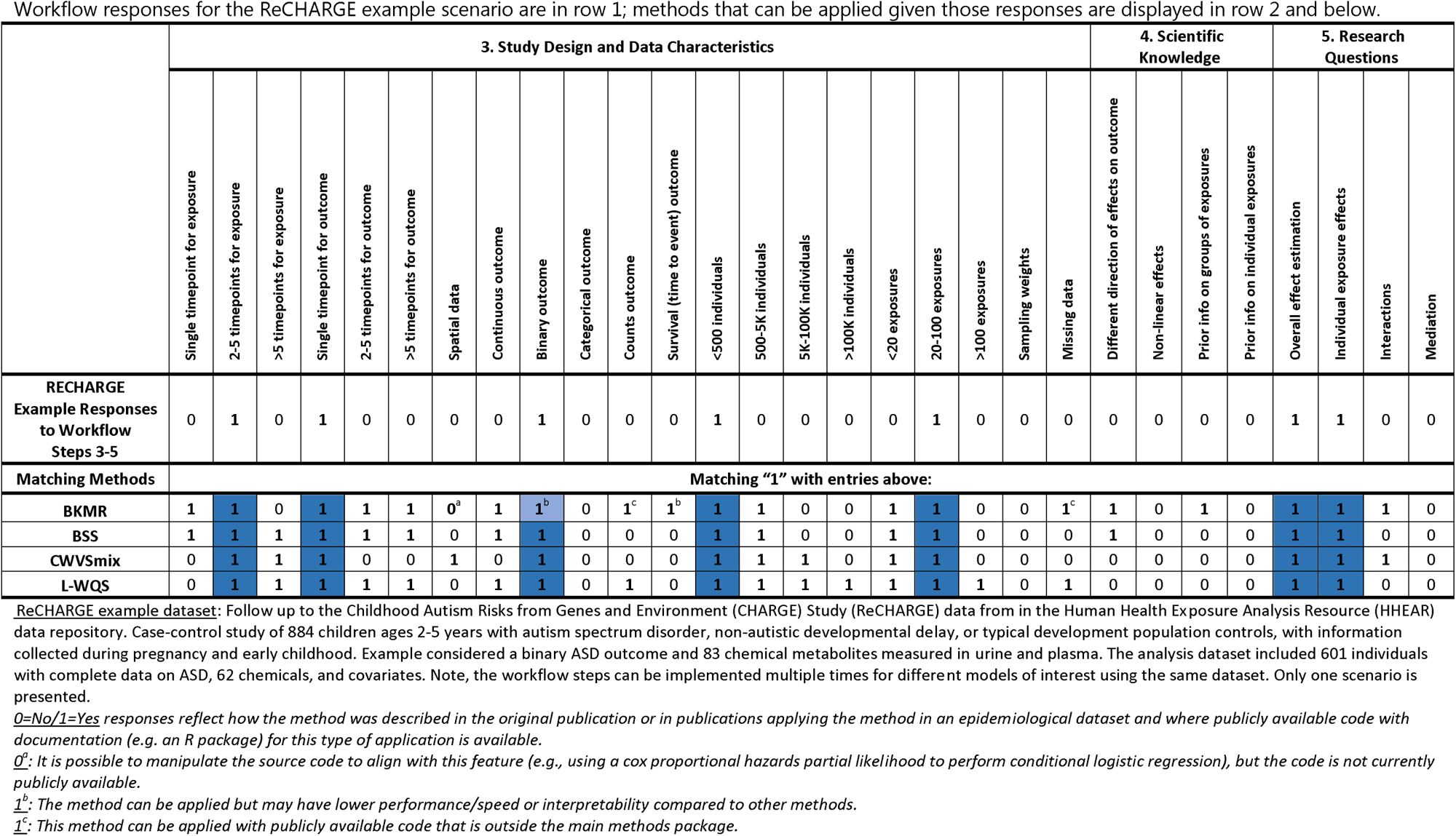
Application of the E-MIX Workflow Steps 3-5: ReCHARGE Example Data.

## Glossary

### Co-exposure amplification bias

Unmeasured confounding can be problematic in mixtures epidemiology. Models that include many environmental exposures can potentially amplify the amount of bias, known as co-exposure amplification bias, depending on the structure of unmeasured confounding, relative to single exposure models. The amount of bias depending on the correlation among the exposure measures and the strength of the unmeasured confounding.

### Out-of-sample predictive performance

General term for the accuracy of a fitted model’s predictions on data not used to fit the model. Accurate out-of-sample predictions suggest the model has found a true statistical relationship in the data, rather than simply being “overfit” to the observations it was trained on.

### MCMC convergence

A Markov Chain Monte Carlo (MCMC) algorithm for Bayesian model fitting has converged when it is generating samples from the posterior distribution of the model parameters.

### Index model for supervised methods

A multi-exposure index is a weighted sum of exposure variables, meant to represent exposure to the mixture of exposures. In supervised index models, the weights of the index are estimated within a health effects model for an outcome, thereby estimating a weighted average most associated with the outcome. Popular examples of index models for environmental mixtures included weighted quantile sum regression and quantile G-computation, with Bayesian multiple index models being a relatively new class of models.

### Response surface model

Characterizes the exposure-response relationship by a multi-dimensional surface that represents how the mean of an outcome varies for any given value of the exposures. Because this surface is often estimated non-parametrically (that is, with minimal assumptions on the shape or structure of this surface), this class of models often allows for estimation of non-linear and non-additive relationships between exposures and response, thereby allowing for a broad array of exposure-response relationships. Examples in environmental mixtures analyses include multivariate generalized additive models, kernel machine regression, and MixSelect.

### Latent factors (latent variable model)

A latent variable model assumes that the joint distribution of observed variables (sometimes referred to as “manifest variables”) can be represented by a smaller number of latent, unobserved, factors. In environmental mixture studies, a latent variable model typically assumed latent variables that generate the joint distribution of multiple exposures and the outcome. Examples include Factor Analysis for Interactions (FIN) and Bayesian profile regression.

### Distributed lag model

A distributed lag model characterizes how an outcome depends on a sequence of values of an exposure. While original applications in the environmental health sciences focused on time series of outcomes, such as how daily mortality counts are associated with daily air pollution exposures up to two weeks (say) prior to the date of death, this class of models has also be popular in children’s health studies focusing on developmental windows of susceptibility to an environmental exposure during pregnancy and childhood. In this context, distributed lag models for environmental mixtures include BKMR-DLM, structured Bayesian regression tree pairs, and multiple exposure distributed lag models with variable selection.

## Acknowledgements

We recognize important contributions to the overall NIEHS Powering Research through Innovative Methods for mixtures in Epidemiology (PRIME) program and to mixtures methods development from NIEHS staff Claudia Thompson, Toccara Chamberlain, Abee Boyles, and David Balshaw and from additional PRIME principal investigators Tom Webster, Chris Gennings, Hua Yun Chen, Mary Turyk, Marie Lynn Miranda, and Kathy Ensor. We sincerely thank the NIEHS Office of Communications and Public Liaison and Web Design Team, including Cheryl Thompson, Stephanie Bishop, Claus Jensen, Qasim Rasheed, David Christie, Joseph Poccia, and Carol Kelly, for their rapid development and ongoing support of the NIEHS Environmental Mixtures Methods Repository.

## Conflict of Interest

The authors have no conflicts of interest to disclose. Example Data

The original NHANES data used in this exercise and example code for data formatting and some methods is available in the supplemental material of Gibson et al., 2019 (https://github.com/lizzyagibson/Mixtures.Workshop.2018). The original RECHARGE data was obtained from the publicly available data in the Human Health Exposure Resource (HHEAR) Data Repository under CHEAR project # 2016-1461; doi 10.36043/1461_222, 10.36043/1461_219, 10.36043/1461_630_2022.2. from the HHEAR Data Center: https://hheardatacenter.mssm.edu/.

