## Supplementary File 1 for "Environmental Mixtures Analysis (E-MIX) Workflow and Methods Repository"

Research

- Scientific Data
- Environmental Mixtures Analysis (E-MIX) Repository
  - Describe Your Data
  - Search Results

### Environmental Mixtures Analysis (E-MIX) Repository

Human exposure to complex, changing, and variably correlated mixtures of environmental chemicals has presented analytical challenges to epidemiologists and human health researchers. There have been wide variety of recent advances in statistical methods for analyzing mixtures data, with most of these methods having open-source software for implementation. This **Environmental Mixtures Methods (E-MIX) Repository** includes information on statistical methods for examining the associations between a mixture of environmental exposures (multiple exposures) and health outcomes, with publicly available software and documentation to guide application.

#### Which Method Should I Use?

Identifying the most appropriate method for a given application can be challenging, particularly for researchers new to environmental mixtures analysis. The **E-MIX Workflow** offers an organized series of steps for approaching environmental mixtures analysis in epidemiological data. The steps in the workflow are intended to be followed sequentially, starting with the overall conceptual model for the analysis of interest (**Step 1**) and data processing and exploratory analyses (**Step 2**). Responses to prompts of the **Steps 3 through 5** can be incorporated in an advanced search of the **E-MIX Repository** in any order and will provide a list of relevant statistical models that can be applied to address a specific context. The final **Step 6** includes considerations for model assessment and evaluation.

**E-MIX Workflow Steps 1 -6**

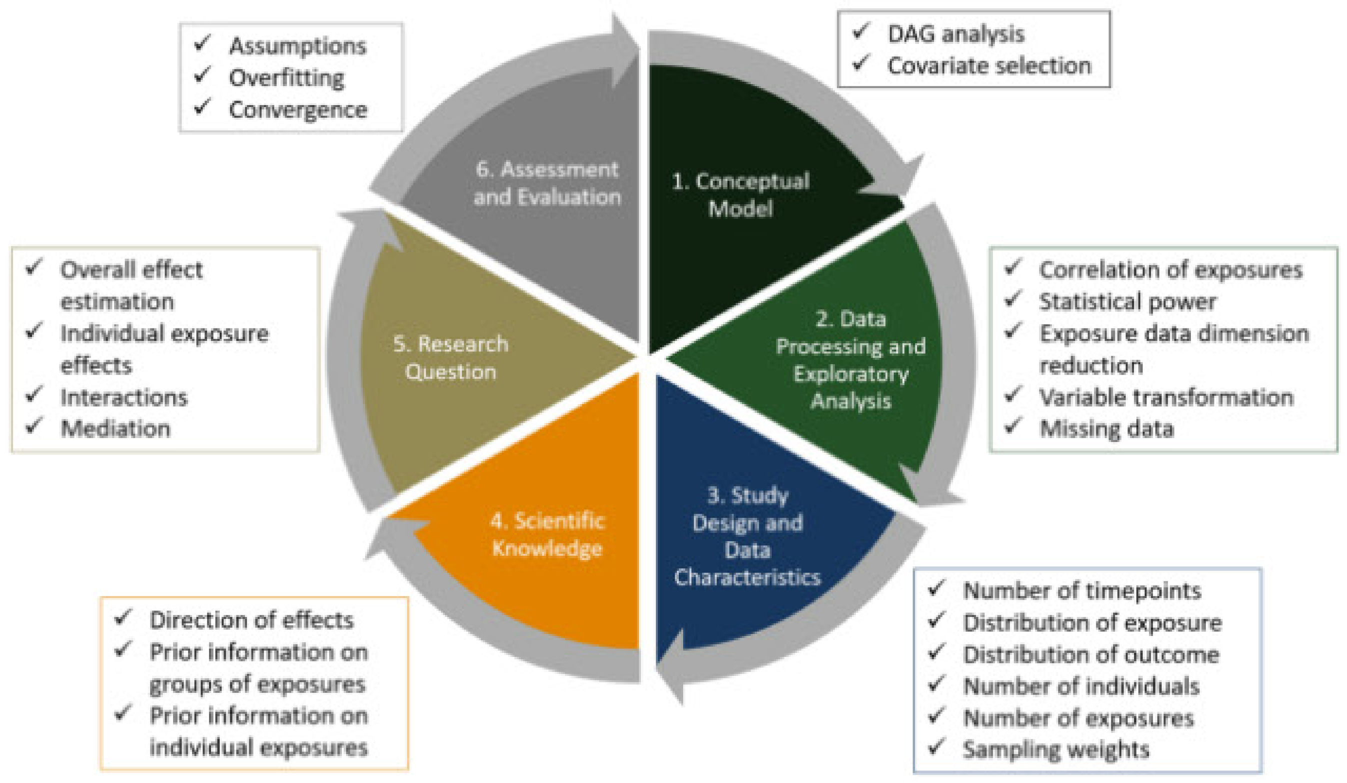


#### Search the Methods

Search

**OR**

Answer questions about your data to find the methods that can be best applied for your research.

Enter Your Data

#### Example Searches

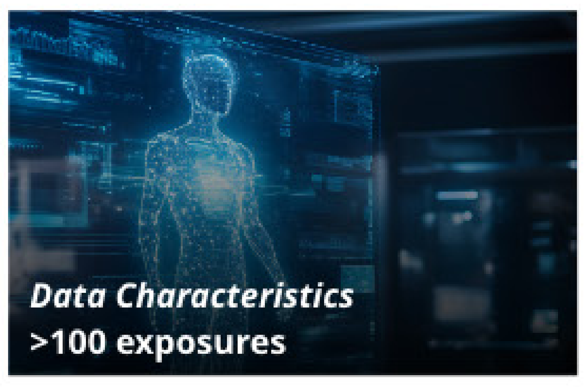


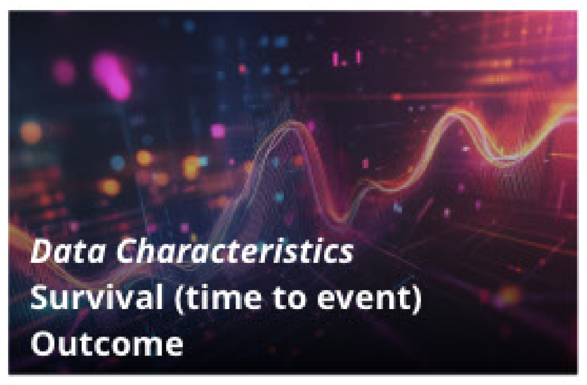


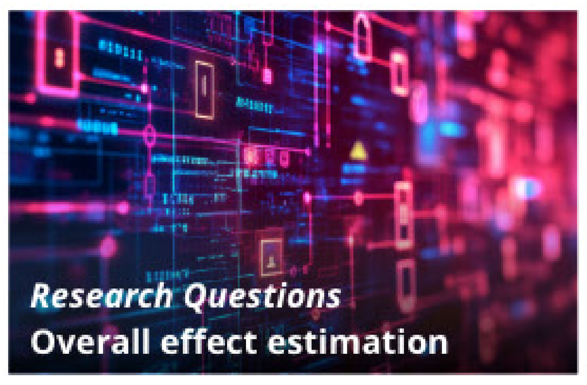


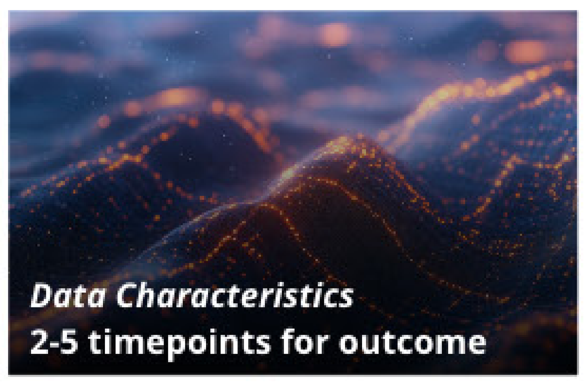


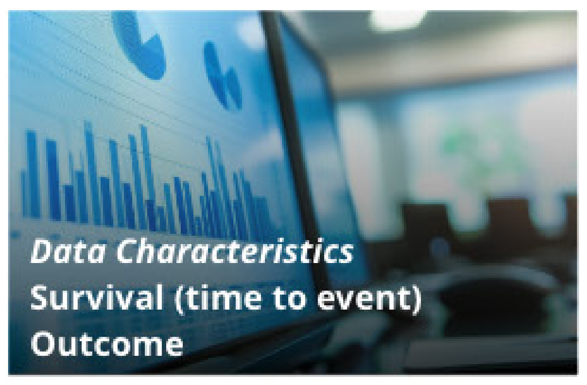


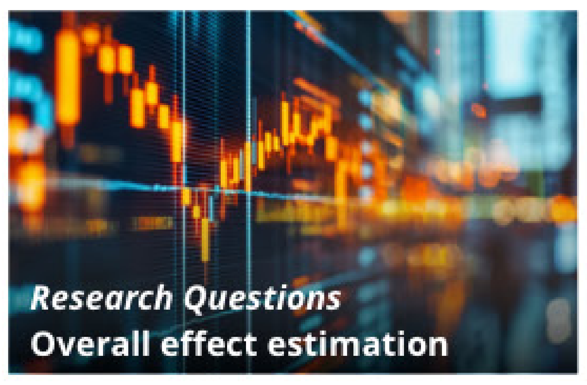

Research

- Scientific Data
- Environmental Mixtures Analysis (E-MIX) Repository
  - Describe Your Data
  - Search Results

### Describe Your Data

#### Environmental Mixtures Analysis (E-MIX) Repository

Study Design and Data Characteristics | Scientific Knowledge | Research Questions

##### Study Design and Data Characteristics

**Timepoint for exposure**

- ○ Single timepoint for exposure
- ○ 2-5 timepoints for exposure
- ○ >5 timepoints for exposure

**Timepoint for outcome**

- ○ Single timepoint for outcome
- ○ 2-5 timepoints for outcome
- ○ >5 timepoints for outcome

**Does your data include spatial data**

- ○ Include spatial data

**Describe the outcome**

- ○ Continuous outcome
- ○ Binary outcome
- ○ Categorical outcome
- ○ Counts outcome

**Survival (time to event) outcome**

- ○ Yes

**Study Size**

- ○ <500 individuals
- ○ 500-5K individuals
- ○ 5K-100K individuals
- ○ >100K individuals

**Number of Exposure**

- ○ <20 exposures
- ○ 20-100 exposures
- ○ >100 exposures

**Sampling weights**

- ○ Yes

**Missing data**

- ○ Yes

Next: Scientific Knowledge

Submit Selections

Research

Scientific Data

Environmental Mixtures Analysis (E-MIX) Repository

Describe Your Data

Search Results

### Describe Your Data

#### Environmental Mixtures Analysis (E-MIX) Repository

Study Design and Data Characteristics | **Scientific Knowledge** | Research Questions

##### Scientific Knowledge

**Different direction of effects on outcome**

- [ ] Yes

**Non-linear effects**

- [ ] Yes

**Prior information on groups of exposures**

- [ ] Yes

**Prior information on individual exposures**

- [ ] Yes

Back: Study Design | Next: Research Questions

Submit Selections

Research

Scientific Data
Environmental Mixtures Analysis (E-MIX) Repository
Describe Your Data
Search Results

### Describe Your Data

#### Environmental Mixtures Analysis (E-MIX) Repository

Study Design and Data Characteristics | Scientific Knowledge | **Research Questions**

##### Research Questions

**Overall effect estimation**

◯ Yes

**Individual exposure effects**

◯ Yes

**Interactions**

◯ Yes

**Mediation**

◯ Yes

Back:
Scientific Knowledge

Submit Selections

### Environmental Mixtures Analysis (E-MIX) Repository

#### Search Results

Search the Repository

##### Refine Your Search

**Study Design and Data Characteristics**

**Timepoint for exposure**
- ☐ Single timepoint
- ☐ 2-5 timepoints
- ☐ >5

**Timepoint for outcome**
- ☐ Single timepoint
- ☐ 2-5 timepoints
- ☐ >5

**Does your data include spatial data**
- ☐ Available

**Describe the outcome**
- ☐ Continuous
- ☐ Binary
- ☐ Categorical
- ☐ Counts

**Survival (time to event) outcome**
- ☐ Yes

**Study Size**
- ☐ <500 individuals
- ☐ 500-5K individuals
- ☐ 5K-100K individuals
- ☐ >100K individuals

**Number of Exposure**
- ☐ <20 exposures
- ☐ 20 - 100 exposures
- ☐ >100 exposures

**Sampling weights**
- ☐ Yes

**Missing data**
- ☐ Yes

Scientific Knowledge

Research Questions

Search Term: **Mixture** Items: **22**

##### Bayes Tree Pairs (BayesTrees)

Considers a situation like BKMR-DLM but uses tree-based models to identify critical windows of exposure for a mixture. Scalable to larger datasets as compared to BKMR-DLM.

##### Bayesian Kernel Machine Regression (BKMR)

Models a real-valued or binary outcome on a set of exposures using a potentially nonlinear exposure-response function modeled by a kernel function. Can accommodate repeated outcome measures using random subject-specific effects.

##### BKMR-Causal Mediation Analysis (BKMR-CMA)

Performs a causal mediation analysis when interest focuses how the effect of a exposure mixture on a real-valued outcome is mediated through a real-valued mediator

##### BKMR-Distributed Lag Model (BKMR-DLM)

Applies when interest focuses on identifying critical windows of exposure for each of multiple exposures within a mixture on a real-valued outcome, and repeated data on each exposure are available on a regular grid (such as weekly during pregnancy). Uses a distributed lag model (DLM) formulation within a Bayesian kernel machine formulation for the exposure.

##### Bayesian Multiple Index Models (BMIM)

Models a real-valued outcome on multiple groups of exposures by assuming group-specific linear combinations of exposures are inputs into a multivariate exposure-response surface.

##### Bayesian Profile Regression (BPR)

Uses a profile formed from a sequence of covariate values, clustered into groups and associated via a regression model to a relevant outcome.

##### Bayesian Subset Selection (BSS)

Can select variables and provide uncertainty quantification for any linear Bayesian model Identify variables to include in subsequent steps

##### Critical Window Variable Selection for Mixtures (CWVSmix)

An extension of the "Critical Window Variable Selection" model of Warren et al. (2020)36 to the mixtures setting.

##### Environmental mixtures with FDR control (EnvMixturesFDR)

Simultaneously estimates the health effects of environmental mixtures and identify important exposures and interactions while controlling FDR.

##### Environmental Risk Score (ERS)

Integrates information on the individual exposure-health outcome effects for multiple exposures by estimating a combined risk score, accounting for covariates, on a portion of the data and then tests for an association between this risk score and the outcome in the remaining data.

1 2 ... 3 Next ›

### Critical Window Variable Selection for Mixtures (CWVSmix)

#### Environmental Mixtures Analysis (E-MIX) Repository

Research
Scientific Data
Environmental Mixtures Analysis (E-MIX) Repository
Resource Details

Return to Search Results

CWVSmix is an extension of the "Critical Window Variable Selection" model of Warren et al. (2020)36 to the mixtures setting.

**Suitable for big data**

Can be used for small datasets <500 individuals and possibly up to 100,000 individuals.

**Software**

Git Hub - CWVSmix

**Reference:** Warren JL, Chang HH, Warren LK, Strickland MJ, Darrow LA, Mulholland JA. Critical Window Variable Selection for Mixtures: Estimating the Impact of Multiple Air Pollutants on Stillbirth. Ann Appl Stat. Sep 2022;16(3):1633-1652. doi:10.1214/21-aoas1560

**Contact:** Joshua Warren

##### Study Design and Data Characteristics

*The model is suitable for the following study design/data contexts and effects of interest.*

**Timepoint(s) for exposure:** Multiple timepoints

**Timepoint(s) for outcome:** Single timepoint

**Distribution of outcome:** Spatial data
Binary outcome

**Number of individuals:** <500
500-5k

**Number of exosures:** <20
20-100
>100 exposures

##### Scientific Knowledge

**Different direction of effects on outcome:** Yes

**Prior information on groups of exposures:** Yes

**Prior infomation on individual exposures:** No

##### Research Questions

**Overall effect estimation:** Yes

**Individual exposure effects** Yes

**Interactions:** Yes

##### Assessment and Evaluation

**Model-specific assumptions:** The model assumes the log odds of the outcome is well-modeled as a linear function of covariates and time-varying exposures with interaction effects. It also assumes that at a given time, all main and interaction coefficients for the exposures have the same sign.

**Risks of overfitting:** Traditional cross-validation can be employed to guard against overfitting by comparing the magnitude of training and testing errors.

**Convergence challenges:** CWVSmix employs Markov Chain Monte Carlo for Bayesian model fitting. Traditional convergence diagnostics (visual inspection of trace plots, Gelman-Rubin statistics applied to multiple chains) should employed as a matter of practice to ensure convergence.
